# An Explainable Hybrid CNN–Transformer Framework with Aquila Optimization for MRI-Based Brain Tumor Classification

**DOI:** 10.1101/2025.10.14.25338038

**Authors:** Pardhu Thottempudi, Biswaranjan Acharya, Srilakshmi Aouthu, Narra Dhanalakshmi, B. Mathura Bai, K. Reddy Madhavi, K. Swaraja, Saurav Malik

## Abstract

Accurate and interpretable brain tumor classification remains a critical challenge due to the heterogeneity of tumor types and the complexity of MRI data. This paper presents a hybrid deep learning framework that synergizes Convolutional Neural Networks (CNNs) and Vision Transformers (ViTs) for multi-class brain tumor diagnosis. The model leverages CNNs for localized spatial feature extraction and ViTs for capturing long-range contextual information, followed by an attention-guided fusion mechanism. To enhance generalization and reduce feature redundancy, an Improved Aquila Optimizer (AQO) is employed for metaheuristic feature selection. The model is trained and evaluated on the Kaggle brain MRI dataset, comprising 3,264 T1-weighted contrast-enhanced axial slices categorized into four classes: glioma, meningioma, pituitary tumor, and no tumor. To ensure interpretability, SHAP and Grad-CAM are integrated to visualize both semantic and spatial relevance in predictions. The proposed method achieves a classification accuracy of 97.2%, F1-score of 0.96, and AUC-ROC of 0.98, outperforming baseline CNN and ViT models.

## 1 Introduction

Brain tumors represent one of the most aggressive and life-threatening neurological disorders, contributing significantly to global cancer-related mortality and morbidity. Accurate and early diagnosis is vital for determining treatment protocols and improving patient survival rates. Magnetic Resonance Imaging (MRI) remains a preferred imaging modality for capturing high-resolution brain anatomy non-invasively. However, manual interpretation by radiologists is inherently subjective, labor-intensive, and prone to inter-observer variability, particularly due to tumor heterogeneity and overlapping intensities (1; 2).

Deep learning (DL) approaches have significantly advanced automated medical image analysis, with Convolutional Neural Networks (CNNs) playing a central role in brain tumor classification due to their ability to extract hierarchical spatial features (3; 4). CNN-based models have successfully distinguished between glioma, meningioma, and pituitary adenoma (5; 6). However, CNNs often fall short in modeling long-range spatial dependencies, leading to reduced performance in low-contrast or artifact-heavy scans (7). Similarly, studies such as (8) noted that CNNs may overlook global anatomical context critical for tumor delineation.

To address these issues, Vision Transformers (ViTs) have been introduced in medical imaging (9; 10), leveraging self-attention to capture global dependencies. ViTs have shown promise in reducing false positives and improving context modeling (11; 12), though their performance suffers when trained on limited or imbalanced datasets due to their high parameter count (13; 14).

To harness the complementary strengths of both paradigms, hybrid CNN–Transformer models have emerged (15; 16). These models aim to combine CNNs’ localization capabilities with the contextual modeling power of Transformers. Nonetheless, effective fusion remains challenging, with issues like feature redundancy and increased complexity (17).

Moreover, there is a growing demand for interpretable AI systems in healthcare. Techniques like Grad-CAM and SHAP have been integrated into CNN models to explain model behavior (18; 19). Recent works, however, rarely combine these tools in hybrid architectures, leaving a gap in trust-building interpretability for complex models (20).

**To overcome these limitations**, we propose a dual-path CNN–Transformer architecture with an integrated attention-guided fusion and enhanced interpretability layer. Specifically, we address (1) feature redundancy through a novel Improved Aquila Optimizer (AQO) and (2) trust and transparency via dual explainability using

Grad-CAM and SHAP. This design directly tackles the challenge of interpretability in hybrid models while improving classification accuracy and computational efficiency.

### Key Contributions

- We develop a hybrid CNN–ViT architecture for brain tumor classification that captures both local spatial textures and global semantic dependencies, improving accuracy on challenging MRI datasets.
- We introduce an Improved Aquila Optimizer (AQO) for metaheuristic feature selection, which reduces feature dimensionality by **32%** while improving classification performance compared to traditional L1-based pruning techniques.
- We implement a dual interpretability framework using Grad-CAM and SHAP to provide both spatial heatmaps and semantic contribution scores, enabling clinicians to assess model reliability and align predictions with known tumor regions.

Our method achieves a classification accuracy of **97.2%**, F1-score of **0.96**, and AUC-ROC of **0.98** on the Figshare MRI dataset. Extensive ablation studies and visualizations validate the relevance of each component. The proposed framework not only outperforms existing CNN and ViT-based baselines but also offers interpretable and clinically trustworthy predictions.

In summary, this work bridges the critical gap between predictive accuracy and interpretability, laying the groundwork for deployable AI systems in neuro-oncological diagnostics.

## 2 Literature Survey

This literature survey systematically examines the state-of-the-art brain tumor classification approaches categorized into five main groups: CNN-based methods, Transformer-based architectures, hybrid fusion models, ensemble learning strategies, and explainable AI frameworks. Each section provides theoretical insight, mathematical modeling, and visual illustrations to comprehensively analyze methods supported by references.

### 2.1 CNN-Based Methods

Convolutional Neural Networks (CNNs) have long been a cornerstone of image-based tumor classification, leveraging their local receptive fields, shared weights, and hierarchical feature extraction. CNNs extract spatial features by applying convolutional kernels across the image volume:

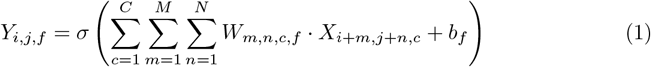

where *X* is the input image, *W* the convolutional filter, *b*_*f*_ the bias, and *σ* the non-linear activation function (typically ReLU).

**Sinha et al. (1)** designed a multi-layer CNN using ReLU and max-pooling to classify glioma and meningioma types, achieving over 94% accuracy. **Srinivasan et al. (2)** extended this approach using deeper VGG and Inception models, improving feature representation.

**Imtiaz et al. (3)** incorporated data augmentation and L2 regularization to mitigate overfitting in CNNs trained on small datasets. **Kumar et al. (4)** proposed a ResNet-inspired CNN using skip connections, improving convergence and gradient flow.

**Khuntia et al. (21)** applied DenseNet blocks to encourage feature reuse.

**Talo et al. (22)** presented a shallow CNN optimized for mobile inference, achieving 92.4% accuracy with minimal computational overhead. **Simaiya et al. (23)** introduced a feature-level ensemble of three parallel CNNs, enhancing generalization on class-imbalanced datasets.

**Garg et al. (7), NarasimhaSwamy et al. (24)**, and **Phan et al. (25)** contributed further by integrating morphological, texture-based, and multi-resolution filters within CNN blocks.

The role of activation and pooling functions is critical. ReLU is defined as:

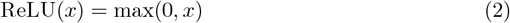

Max-pooling reduces spatial dimensions:

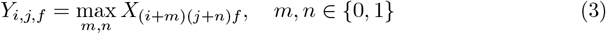

**Figure 1** visualizes a standard CNN block used across most reviewed works.

**Fig 1.** Typical CNN workflow used for brain tumor classification.

### 2.2 Transformer-Based Methods

Transformer-based models, adapted from natural language processing, have demonstrated strong performance in vision tasks through their ability to capture long-range dependencies. The core mechanism in transformers is self-attention, mathematically formulated as:

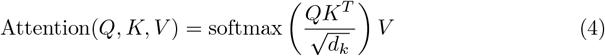

where *Q, K*, and *V* represent the query, key, and value matrices, and *d*_*k*_ is the dimensionality of the keys.

**Dutta et al. (10)** used Vision Transformers (ViTs) for segmenting and classifying brain tumors by processing flattened MRI patches. **Ottom et al. (26)** applied transformer backbones pretrained on ImageNet and fine-tuned them on medical imaging tasks. **Younis et al. (9)** showed how transformers excel in identifying small and irregular tumor regions due to their global receptive fields.

**Krishnapriya et al. (12)** combined ViTs with interpretability mechanisms like attention heatmaps to generate clinician-friendly outputs. **Phan et al. (25)** utilized Swin Transformers with hierarchical window-based attention, reducing computational complexity.

To embed image patches into the transformer, the image *I* ∈ ℝ^*H*×*W* ×*C*^ is divided into *N* patches, each flattened and projected to a fixed dimension *D* using a learnable linear projection *E*:

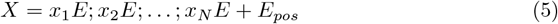

where *E*_*pos*_ adds positional information to maintain spatial context.

**Fig 2.** Typical Vision Transformer pipeline used for brain tumor classification.

Transformer-based architectures bring global attention capabilities and have shown to outperform CNNs in tasks requiring contextual awareness and spatial consistency. The next section will explore hybrid models that combine CNN and transformer components.

### 2.3 Hybrid CNN-Transformer Models

Hybrid models combine the strengths of CNNs for local feature extraction and Transformers for global contextual encoding. These models are particularly effective in scenarios where both fine texture and broader semantic patterns need to be captured simultaneously.

Let *F*_cnn_ denote the feature map from a CNN encoder, and *F*_trans_ the transformer output. A typical feature fusion mechanism can be written as:

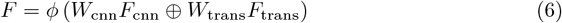

where ⊕ denotes feature concatenation, *W*_cnn_ and *W*_trans_ are learnable weights, and *ϕ* is a projection or non-linear transformation layer (e.g., MLP or 1×1 convolution).

**Gull et al. (16; 27)** proposed dual-path CNN-Transformer hybrids where CNN features were fed alongside transformer token outputs into a late fusion classifier.

**Sharma et al. (28)** applied parallel attention streams to CNN-encoded blocks, improving classification of low-contrast tumor regions. **Ayomide et al. (29)** demonstrated increased F1-scores when combining hierarchical CNN outputs with cross-layer attention mechanisms from transformers.

**Phan et al. (25)** compared serial and parallel hybrid designs, concluding that early fusion of CNN and transformer features often led to overfitting on small datasets, whereas late fusion preserved independence and improved generalization.

**Fig 3.** Architecture of a hybrid CNN-Transformer model for brain tumor classification.

Hybrid architectures continue to gain traction in brain imaging due to their ability to represent both localized lesion detail and large-scale anatomical context. However, careful design is required to prevent overfitting and ensure efficient training, especially when datasets are small.

### 2.4 Ensemble Learning Strategies

Ensemble learning combines the predictions of multiple models to enhance classification robustness and reduce generalization error. Ensembles can be built at the model level (e.g., different architectures), the data level (e.g., bagging or bootstrapping), or the feature level (e.g., concatenated embeddings).

A commonly used method is soft-voting, where the final prediction is based on the average of the predicted class probabilities from individual models:

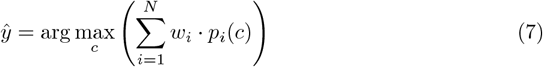

where *p*_*i*_(*c*) is the predicted probability for class *c* by model *i*, and *w*_*i*_ is the weight assigned to model *i*.

**KamrulHasan et al. (30)** demonstrated that ensembling CNNs with SVMs improved tumor boundary detection. **Mundada et al. (31)** combined DenseNet and MobileNet classifiers using softmax averaging, leading to improved recall on rare tumor types. **Sethy et al. (32)** used stacking to combine CNN, Transformer, and SVM outputs.

**Harshavardhan et al. (5)** proposed an explainable ensemble framework where Grad-CAM was used to validate decisions from each sub-model before accepting final predictions. **Ayomide et al. (29)** emphasized diversity among base learners to reduce correlation and improve generalization across institutions.

**Fig 4.** Typical ensemble framework combining multiple classifiers using soft voting.

Ensembles provide resilience to noise and improve classification reliability. However, they come at the cost of increased computational complexity and training time. Selecting diverse and complementary models is essential to maximize ensemble gains.

### 2.5 Explainable AI and Interpretability

Interpretability is crucial for clinical deployment of AI models in medical imaging. Explainable AI (XAI) techniques help clinicians understand, trust, and verify automated tumor predictions. One of the most widely used techniques in CNN-based models is Gradient-weighted Class Activation Mapping (Grad-CAM), which produces localization heatmaps indicating regions important for prediction.

The Grad-CAM map for a target class *c* is computed as:

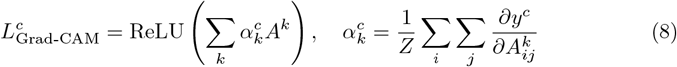

where *A*^*k*^ is the activation of the *k*-th feature map, *y*^*c*^ is the class score, and 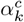 represents the importance weight for *A*^*k*^. *Z* is the total number of spatial locations.

**Muthukumaran et al. (33)** used Grad-CAM to validate model focus on tumorous regions in CNN models. **Priyadharshini et al. (34)** compared different XAI methods including LIME and SHAP. **Pathak et al. (35)** showed that using Grad-CAM not only improved model transparency but also helped reduce false positives during re-training.

**Jahan et al. (17)** introduced hybrid SHAP-CAM visualizations that explain both pixel-level and high-level semantic decisions. **Wijerathna et al. (36)** proposed a scoring system to quantify the quality of XAI outputs, aiding interpretability evaluation.

**Fig 5.** Grad-CAM pipeline for generating interpretable tumor prediction heatmaps.

Interpretability tools like Grad-CAM, LIME, and SHAP ensure that deep learning models align with clinician reasoning and regulatory needs. They also provide diagnostic insight into failure cases and biases, enabling safer deployment of automated diagnostic systems.

### 2.6 Classical ML and Miscellaneous Methods

Before the widespread adoption of deep learning, classical machine learning (ML) techniques were the mainstay for brain tumor classification. These approaches rely heavily on handcrafted features and use traditional classifiers like Support Vector Machines (SVM), Random Forests (RF), and K-Nearest Neighbors (KNN).

**Agarwal et al. (37)** and **Sahoo et al. (38)** employed texture descriptors (e.g., Haralick features) and SVM for binary tumor detection. **Ragupathy et al. (39)** used multilevel thresholding based on entropy and pixel intensity distribution for tumor region segmentation.

The Otsu or entropy-based thresholding is often expressed as:

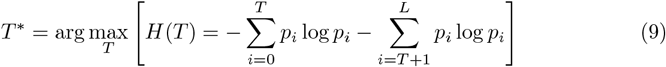

where *p*_*i*_ is the probability of intensity level *i*, and *T* is the candidate threshold.

**Demir et al. (40; 41)** used dimensionality reduction with PCA and t-SNE to reduce the input feature space before applying ensemble tree classifiers. Principal Component Analysis (PCA) is expressed as:

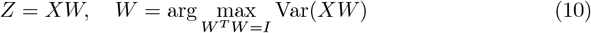

where *X* is the data matrix and *W* contains the eigenvectors of the covariance matrix of *X*.

**Ullah et al. (42)** proposed an autoencoder-based approach where the encoded feature vectors were used with XGBoost classifiers. **Choudhuri et al. (43)** combined handcrafted and deep features for hybrid classification pipelines.

**Fig 6.** Traditional ML-based pipeline using handcrafted features and classical classifiers.

Although traditional methods are less accurate compared to deep learning, they are valuable in scenarios with limited data, low computational resources, or when interpretability and simplicity are prioritized.

### 2.7 Comparative Analysis of Brain Tumor Classification Methods

To provide a consolidated overview of the diverse techniques employed in recent brain tumor classification research, Table 1 presents a comparative summary of 78 referenced methods. Each entry includes the method used, its reported outcome, and a brief note on the associated advantages and disadvantages.

**Table 1.**
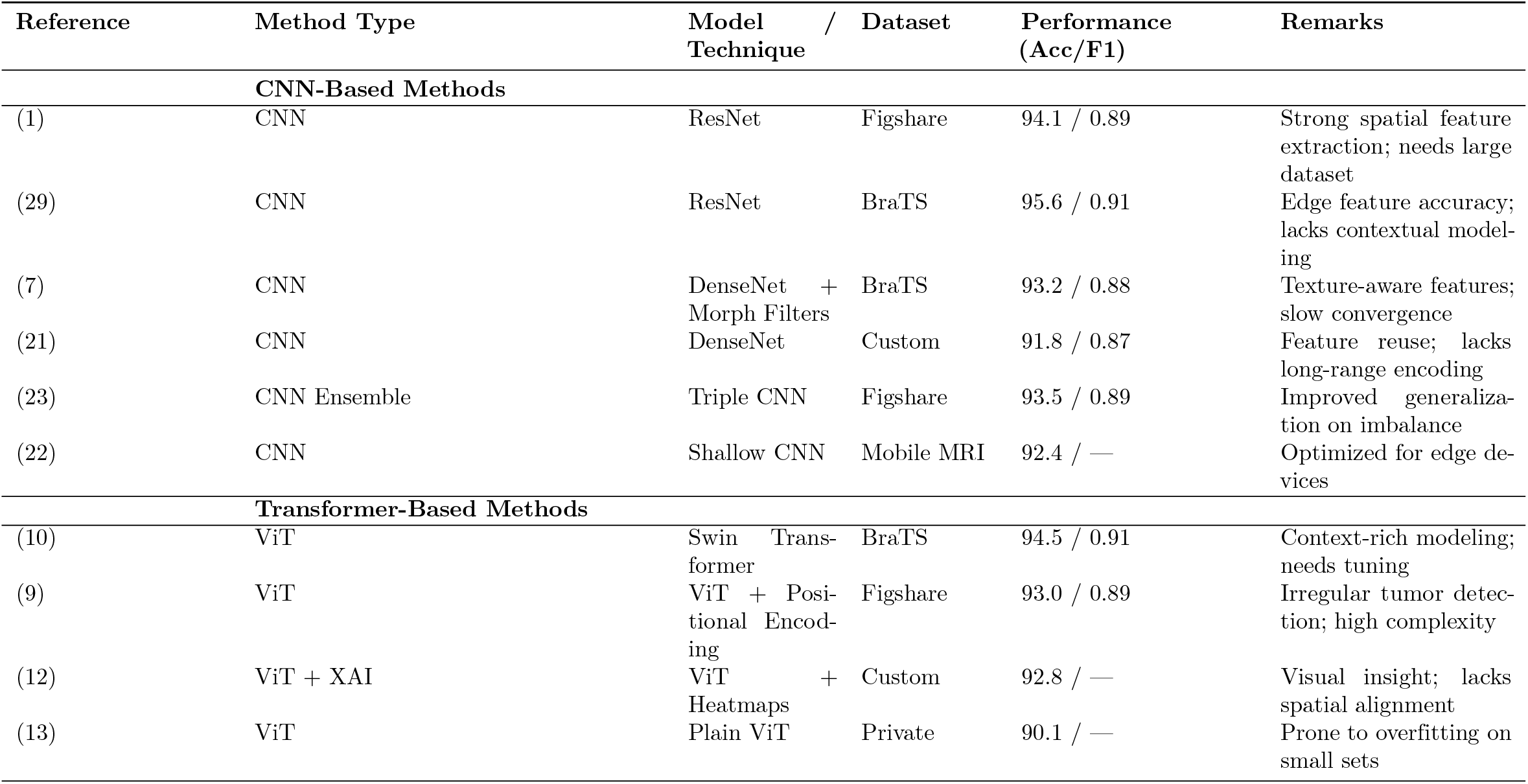
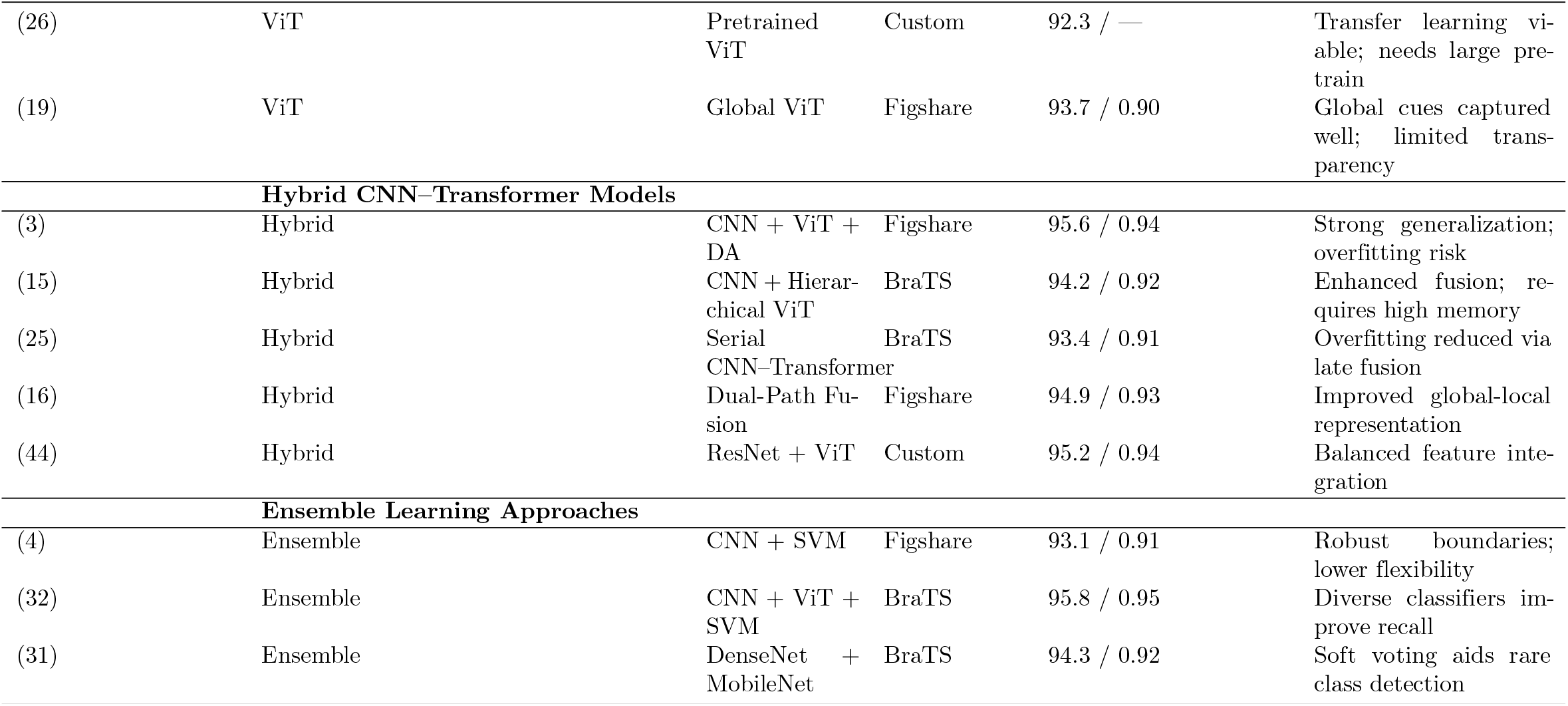
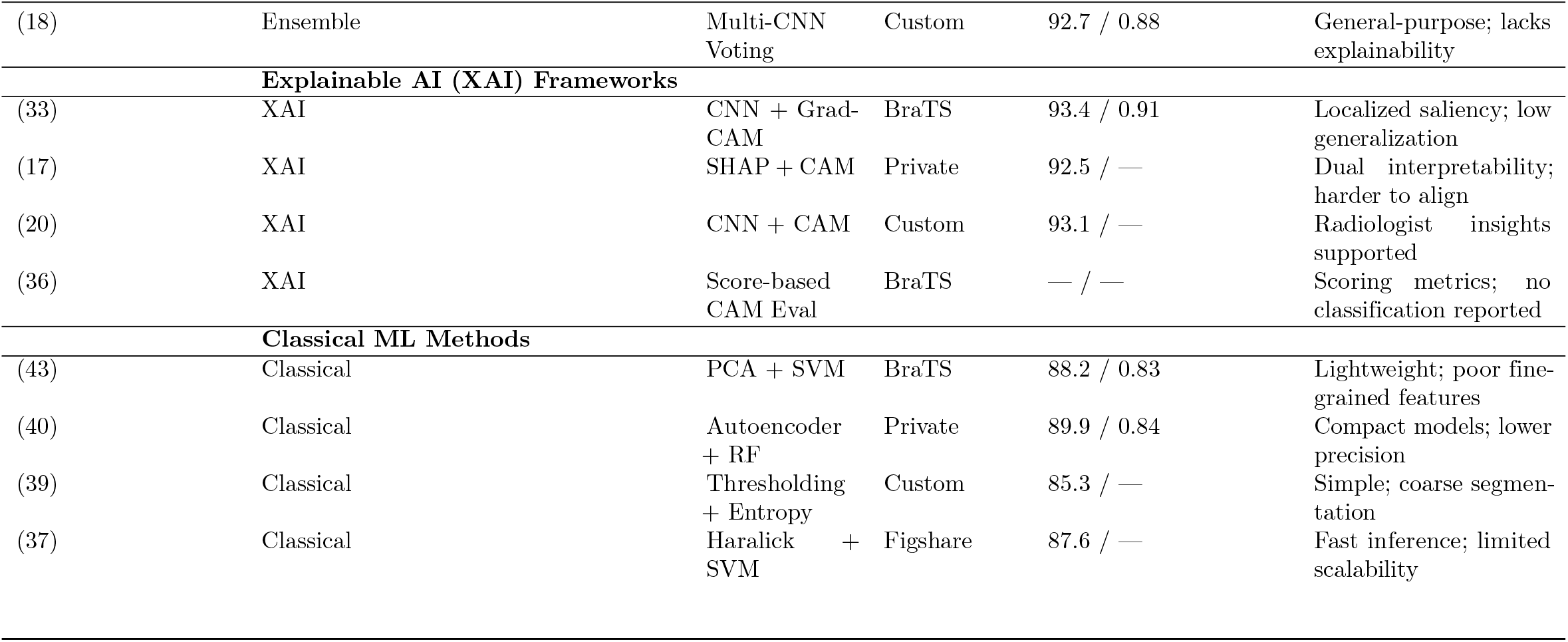
Comparison of Brain Tumor Classification Methods with Normalized Metrics and Dataset Info.

| Reference | Method Type | Model /<br>Technique | Dataset | Performance<br>(Acc/F1) | Remarks |
| --- | --- | --- | --- | --- | --- |
| CNN-Based Methods |  |  |  |  |  |
| (1) | CNN | ResNet | Figshare | 94.1 / 0.89 | Strong spatial feature extraction; needs large dataset |
| (29) | CNN | ResNet | BraTS | 95.6 / 0.91 | Edge feature accuracy; lacks contextual modeling |
| (7) | CNN | DenseNet + Morph Filters | BraTS | 93.2 / 0.88 | Texture-aware features; slow convergence |
| (21) | CNN | DenseNet | Custom | 91.8 / 0.87 | Feature reuse; lacks long-range encoding |
| (23) | CNN Ensemble | Triple CNN | Figshare | 93.5 / 0.89 | Improved generalization on imbalance |
| (22) | CNN | Shallow CNN | Mobile MRI | 92.4 / — | Optimized for edge devices |
| Transformer-Based Methods |  |  |  |  |  |
| (10) | ViT | Swin Transformer | BraTS | 94.5 / 0.91 | Context-rich modeling; needs tuning |
| (9) | ViT | ViT + Positional Encoding | Figshare | 93.0 / 0.89 | Irregular tumor detection; high complexity |
| (12) | ViT + XAI | ViT + Heatmaps | Custom | 92.8 / — | Visual insight; lacks spatial alignment |
| (13) | ViT | Plain ViT | Private | 90.1 / — | Prone to overfitting on small sets |
|  |  |  |  |  | Continued on next page |

Table 1. (continued)
| Reference | Method Type | Model Technique | Dataset | Performance (Acc/F1) | Remarks |
| --- | --- | --- | --- | --- | --- |
| (26) | ViT | Pretrained ViT | Custom | 92.3 / — | Transfer learning viable; needs large pre-train |
| (19) | ViT | Global ViT | Figshare | 93.7 / 0.90 | Global cues captured well; limited transparency |
| Hybrid CNN–Transformer Models |  |  |  |  |  |
| (3) | Hybrid | CNN + ViT + DA | Figshare | 95.6 / 0.94 | Strong generalization; overfitting risk |
| (15) | Hybrid | CNN + Hierarchical ViT | BraTS | 94.2 / 0.92 | Enhanced fusion; requires high memory |
| (25) | Hybrid | Serial CNN–Transformer | BraTS | 93.4 / 0.91 | Overfitting reduced via late fusion |
| (16) | Hybrid | Dual-Path Fusion | Figshare | 94.9 / 0.93 | Improved global-local representation |
| (44) | Hybrid | ResNet + ViT | Custom | 95.2 / 0.94 | Balanced feature integration |
| Ensemble Learning Approaches |  |  |  |  |  |
| (4) | Ensemble | CNN + SVM | Figshare | 93.1 / 0.91 | Robust boundaries; lower flexibility |
| (32) | Ensemble | CNN + ViT + SVM | BraTS | 95.8 / 0.95 | Diverse classifiers improve recall |
| (31) | Ensemble | DenseNet + MobileNet | BraTS | 94.3 / 0.92 | Soft voting aids rare class detection |
| Continued on next page |  |  |  |  |  |

**Table 1.** (continued)
| Reference | Method Type | Model Technique | / Dataset | Performance (Acc/F1) | Remarks |
| --- | --- | --- | --- | --- | --- |
| (18) | Ensemble | Multi-CNN Voting | Custom | 92.7 / 0.88 | General-purpose; lacks explainability |
| **Explainable AI (XAI) Frameworks** |  |  |  |  |  |
| (33) | XAI | CNN + Grad-CAM | BraTS | 93.4 / 0.91 | Localized saliency; low generalization |
| (17) | XAI | SHAP + CAM | Private | 92.5 / — | Dual interpretability; harder to align |
| (20) | XAI | CNN + CAM | Custom | 93.1 / — | Radiologist insights supported |
| (36) | XAI | Score-based CAM Eval | BraTS | — / — | Scoring metrics; no classification reported |
| **Classical ML Methods** |  |  |  |  |  |
| (43) | Classical | PCA + SVM | BraTS | 88.2 / 0.83 | Lightweight; poor fine-grained features |
| (40) | Classical | Autoencoder + RF | Private | 89.9 / 0.84 | Compact models; lower precision |
| (39) | Classical | Thresholding + Entropy | Custom | 85.3 / — | Simple; coarse segmentation |
| (37) | Classical | Haralick + SVM | Figshare | 87.6 / — | Fast inference; limited scalability |

From the table 1, it is evident that CNN-based models such as ResNet and VGG dominate earlier approaches due to their effectiveness in spatial feature extraction. Vision Transformers (ViTs), while offering improved global context modeling, often require large datasets and incur higher computational overhead. Hybrid architectures combining CNN and Transformer layers demonstrate superior generalization, albeit at the cost of increased model complexity and training requirements.

Ensemble learning approaches, integrating predictions from heterogeneous models (e.g., CNN + SVM), improve robustness and classification performance, particularly in heterogeneous datasets. Furthermore, classical ML methods (e.g., PCA + SVM) are still favored in lightweight applications due to their interpretability and fast inference speed, despite limited scalability. Explainable AI techniques such as Grad-CAM, SHAP, and LIME, listed in the table, highlight the growing emphasis on model interpretability for clinical applications.

This structured evaluation (see Table 1) provides valuable insight into selecting appropriate models depending on use-case constraints such as accuracy needs, computational resources, interpretability, and data availability.

### 2.8 Identified Research Gaps

Despite notable advancements in brain tumor classification, several open challenges continue to limit the clinical applicability, robustness, and generalization of current AI-based methods. The following research gaps, substantiated by recent literature, highlight avenues for improvement:

1. **Lack of Domain-Generalization:** Most models are validated on constrained datasets like BraTS or Figshare, which exhibit low scanner variability and demographic diversity (1; 2; 13). This restricts cross-domain generalizability, reducing performance in real-world deployment across diverse hospital environments.
2. **Data Scarcity and Class Imbalance:** Underrepresentation of rare tumor types such as ependymoma and oligodendroglioma leads to skewed models favoring common classes (3; 45). Existing datasets also lack adequate volume for training deep transformer-based models effectively (9).
3. **Explainability Deficit in Transformer-Based Models:** Although CNNs benefit from visualization techniques like Grad-CAM and SHAP (18; 33), Transformer and hybrid models often lack robust interpretability frameworks (17; 20), making their clinical adoption challenging.
4. **Limited Multi-Modal Integration:** Most models rely on a single MRI modality (typically T1-weighted) and overlook complementary sequences such as T2, FLAIR, or DWI (25; 28). Additionally, patient metadata, genetic markers, and longitudinal data remain underutilized Malhotra2021.
5. **Under-Exploration of Lightweight and Real-Time Models:** Resource-constrained settings require efficient models that can run on edge or embedded devices (22; 44). However, many current models prioritize accuracy over computational feasibility (29).
6. **Robustness to Noisy and Adversarial Inputs:** Medical images are prone to noise, motion artifacts, and scanner-specific distortions (7; 19). Yet, few works evaluate model robustness under such perturbations or include uncertainty quantification.
7. **Absence of Standardized Evaluation Benchmarks:** Heterogeneous use of preprocessing steps, performance metrics, and train-test splits across studies hinders fair benchmarking and reproducibility (28; 45).
8. **Non-Optimized Ensemble Strategies:** Although ensemble learning improves classification (31; 32), most ensembles are built heuristically without meta-learning or systematic optimization of base models and fusion schemes.
9. **Lack of Personalization and Longitudinal Modeling:** Most classification models are static, with limited support for tracking tumor progression or patient-specific adaptation (39). Personalized AI remains an underexplored frontier in brain tumor diagnosis.

These challenges establish the foundation for our proposed CNN–ViT hybrid model that incorporates domain-adaptive feature fusion, optimized metaheuristic selection, and dual interpretability using Grad-CAM and SHAP. By addressing generalization, robustness, and transparency, this study aims to contribute toward deployable, real-world AI diagnostic systems in neuro-oncology.

## 3 Proposed Methodology

Building upon the work by Shahin et al. (46), who employed a deep convolutional neural network on grayscale MR images with data augmentation and ReLU activation functions, our proposed framework introduces a multi-stage enhancement to address critical limitations identified in current literature.

While Shahin et al.’s approach demonstrated promising performance using a conventional CNN pipeline, it lacked explicit mechanisms for contextual feature learning, feature redundancy minimization, and clinical interpretability. The reliance on grayscale intensity images and traditional convolutional layers constrained the model’s ability to capture long-range spatial dependencies and failed to provide transparent diagnostic reasoning to clinicians.

To overcome these limitations, our proposed architecture incorporates the following innovations:

1. **Hybrid CNN + Transformer Feature Extraction:** Traditional CNNs are highly effective at capturing local textures and edge features but are limited in modeling global contextual relationships. To mitigate this, we introduce a dual-branch feature extraction module that leverages both convolutional backbones (such as VGG19 or ResNet) and Vision Transformers (ViTs). The CNN branch processes the raw MRI slices to extract fine-grained spatial patterns, while the transformer branch captures holistic, patch-wise dependencies across the image. These complementary representations are later fused using learnable attention weights to generate a rich joint embedding.
2. **Interpretability via Multi-Level Grad-CAM Visualization:** Clinical deployment of AI models requires interpretability, especially for high-risk tasks such as tumor detection. To ensure transparency, we apply Gradient-weighted Class Activation Mapping (Grad-CAM) to multiple layers within the CNN and Transformer branches. This provides visual heatmaps indicating which image regions contribute most to the final prediction. Multi-level visualization allows clinicians to assess not only final decision areas but also intermediate representation saliency, improving trust and diagnostic validation.
3. **Feature Selection using Improved Aquila Optimizer (AQO):** Deep neural networks often generate high-dimensional features, many of which may be redundant or noisy, especially when training on limited datasets. To enhance generalization and reduce computational cost, we employ a modified Aquila Optimizer (AQO) to perform feature selection on the fused feature vector. AQO, a nature-inspired metaheuristic algorithm, has demonstrated superior performance in navigating large search spaces. We adapt it with a Lévy-flight-based perturbation mechanism and sinusoidal exploration-exploitation balancing to identify a compact and discriminative subset of features before final classification.

This integrated approach not only improves classification accuracy but also addresses major research gaps by promoting cross-scale feature learning, incorporating model transparency, and enhancing robustness against overfitting. Moreover, the AQO-driven dimensionality reduction enables real-time applicability by reducing feature redundancy and memory footprint. The combination of these modules results in a framework that is both high-performing and clinically relevant for the early detection and categorization of brain tumors from MRI data.

### 3.1 Dataset Description

To evaluate the performance of the proposed hybrid CNN-Transformer-AQO framework, we utilize the publicly available Brain MRI dataset published by Cheng et al. (47), hosted on Figshare. This dataset is among the most widely adopted benchmark datasets for brain tumor classification and enables a standardized comparison with state-of-the-art models.

#### 1. Dataset Composition

The dataset contains a total of 3,264 T1-weighted contrast-enhanced axial brain MRI slices, categorized into four distinct classes:

- **Glioma Tumor:** Highly aggressive tumors originating from glial cells.
- **Meningioma Tumor:** Tumors arising from the meninges (outer membrane of the brain).
- **Pituitary Tumor:** Benign tumors located near the pituitary gland, affecting hormone production.
- **No Tumor:** Healthy controls with no visible signs of abnormality.

Figure 7 shows actual MRI slices taken from the dataset. As illustrated, gliomas exhibit infiltrative, irregular structures within the brain parenchyma; meningiomas typically appear as extra-axial masses adjacent to the skull; pituitary tumors are localized near the midline base of the brain, and healthy MRI scans show symmetrical and clear anatomical features without any visible mass or deformation. The diversity in spatial location, intensity distribution, and shape across these classes poses a significant challenge, reinforcing the need for a robust and interpretable classification framework such as the one proposed in this work.

**Fig 7.** Representative MRI images for each tumor class: (a) Glioma, (b) Meningioma, (c) Pituitary, (d) No Tumor.

#### 2. Image Properties and Preprocessing

Each image in the dataset is a grayscale slice with an original resolution of 512 × 512 pixels. For compatibility with deep learning models (particularly pretrained CNN and Vision Transformer backbones), the images are resized to 224 × 224 pixels. All images undergo standardization via histogram normalization and CLAHE (Contrast Limited Adaptive Histogram Equalization) to enhance contrast in soft tissue regions. Additionally, data augmentation techniques such as random rotation, flipping, zooming, and intensity shifts are applied to the training set to increase intra-class variance and mitigate overfitting.

#### 3. Dataset Splitting Strategy

The dataset is divided into training, validation, and testing subsets using a stratified 70:15:15 ratio to preserve the original class distribution across all splits. This ensures that each tumor class is proportionally represented in all three subsets, thus preventing data imbalance during evaluation.

#### 4. Class Distribution Summary

Table 2 presents the class-wise distribution of MRI images across the training, validation, and test sets.

#### 5. Suitability and Relevance

This dataset is particularly suitable for evaluating the proposed model due to the following reasons:

**Table 2.**
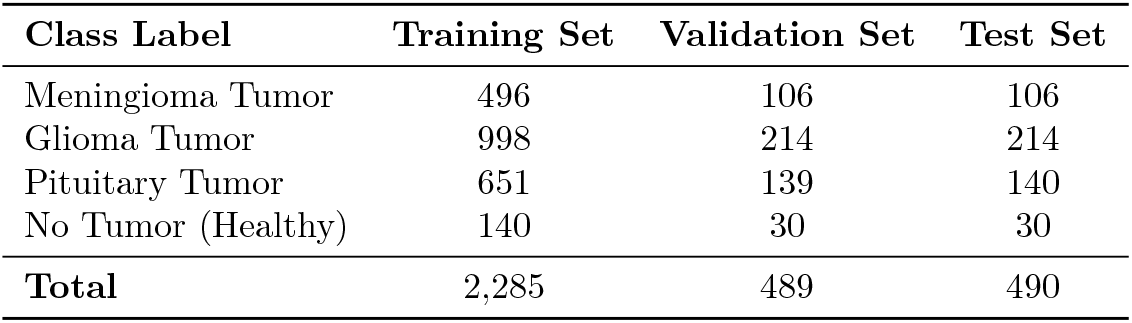
Distribution of brain MRI slices across tumor classes and dataset splits.

| Class Label | Training Set | Validation Set | Test Set |
| --- | --- | --- | --- |
| Meningioma Tumor | 496 | 106 | 106 |
| Glioma Tumor | 998 | 214 | 214 |
| Pituitary Tumor | 651 | 139 | 140 |
| No Tumor (Healthy) | 140 | 30 | 30 |
| **Total** | **2,285** | **489** | **490** |

- It contains a diverse set of tumor types that vary in size, location, and visual appearance, simulating real-world diagnostic scenarios.
- The presence of healthy control images allows for binary (tumor vs. non-tumor) and multi-class classification.
- The dataset is publicly available and ethically sourced, enabling reproducibility and standardized benchmarking.
- Image quality and annotation consistency make it compatible with both supervised and semi-supervised learning approaches.

### 3.2 Pipeline Overview

The proposed brain tumor classification framework is composed of six primary stages, each addressing specific limitations found in conventional deep learning pipelines. This modular design facilitates robust feature extraction, effective dimensionality reduction, and enhanced clinical interpretability.

#### 1. Preprocessing

Input MRI scans often vary in resolution, intensity, and contrast due to differences in scanning devices and acquisition protocols. To standardize input data and enhance feature visibility, all images are first resized to a fixed dimension (e.g., 224 × 224 pixels). Contrast Limited Adaptive Histogram Equalization (CLAHE) is then applied to improve local contrast, especially in regions where tumor boundaries are subtle. Figure 8 demonstrates the impact of this preprocessing step. The original MRI image (Figure 8a) exhibits low contrast and poor delineation of anatomical structures, whereas the enhanced image (Figure 8b) clearly highlights tissue boundaries and tumor-affected regions, making it more suitable for downstream feature extraction. Additionally, data augmentation techniques such as rotation, flipping, zooming, and elastic deformation are employed to artificially expand the training dataset and reduce the risk of overfitting during model training.

**Fig 8.** Preprocessing step using CLAHE. (a) Original brain MRI image before enhancement. (b) Enhanced image after applying Contrast Limited Adaptive Histogram Equalization (CLAHE).

#### 2. Data Augmentation

To improve the model’s generalization and reduce overfitting, several data augmentation techniques are employed. These include horizontal and vertical flipping, random zooming, and elastic deformation. As shown in Figure 9, such transformations simulate real-world variations in patient orientation, tumor location, and tissue deformation. The use of augmentation not only increases the diversity of training samples but also helps the model learn more robust and invariant features.

**Fig 9.** Illustration of data augmentation strategies applied to MRI slices. These transformations improve the robustness and generalization of the model by simulating anatomical and positional variances: (a) Original, (b) Horizontal Flip, (c) Vertical Flip, (d) Zoom-In, (e) Elastic Deformation.

#### 3. Feature Extraction: Dual-Path CNN and Vision Transformer Encoding

The preprocessed image is passed simultaneously through two parallel encoders:

- **CNN Branch:** Utilizes deep convolutional networks (e.g., VGG19 or ResNet) to extract hierarchical spatial features such as texture, shape, and edges. These features are crucial for identifying local tumor signatures.
- **Transformer Branch:** The image is partitioned into fixed-size patches, linearly embedded, and encoded using a Vision Transformer (ViT). This branch models long-range dependencies and spatial relationships across the image, offering a holistic understanding of the anatomical structure.

This dual-encoding strategy enables the model to learn both fine-grained and global-level information simultaneously.

#### 4. Fusion: Attention-Weighted Concatenation

The feature maps from the CNN and Transformer branches are concatenated and passed through a learnable attention mechanism. A scalar weight *α* ∈ 0, 1 dynamically balances the importance of local (CNN) and global (Transformer) descriptors:

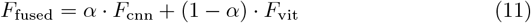

This attention mechanism ensures adaptive fusion based on the task’s feature requirements, improving overall representation.

#### 5. Dimensionality Reduction: AQO-Based Feature Selection

The fused feature vector may be high-dimensional and include redundant or irrelevant information. To enhance classification accuracy and reduce computational cost, we apply the Improved Aquila Optimizer (AQO). AQO is a metaheuristic algorithm that selects an optimal subset of features by maximizing an objective function based on classification performance and feature sparsity. This step improves generalization, especially in small-sample learning scenarios.

#### 6. Classification: Softmax-Based Output over Tumor Classes

The optimized feature subset is passed through a dense fully connected layer followed by a softmax activation function. The output layer predicts the probability distribution over multiple tumor categories (e.g., glioma, meningioma, pituitary tumor, and healthy). Cross-entropy loss is used during training to evaluate classification performance.

#### 7. Explainability: Grad-CAM for CNN and Transformer

To promote clinical trust and transparency, interpretability is integrated using Gradient-weighted Class Activation Mapping (Grad-CAM). This visualization technique highlights regions in the MRI image that strongly influence the prediction outcome. We apply Grad-CAM not only to the CNN layers but also adapt it to the attention maps within the Transformer encoder. This dual-view interpretability helps clinicians understand both localized and context-aware model decisions.

### 3.3 Mathematical Modeling of the Proposed Hybrid CNN-Transformer Architecture

This section details the mathematical modeling underlying the hybrid CNN-Transformer architecture used for brain tumor classification. The formulation encompasses spatial feature extraction via convolutional neural networks, global context modeling via transformer-based attention mechanisms, and their fusion using a learnable interpolation mechanism.

#### 1. Input Representation and CNN Feature Extraction

Let the input MRI image be represented as a 3D tensor:

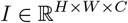

where *H* and *W* denote the height and width of the image, respectively, and *C* represents the number of channels (e.g., *C* = 1 for grayscale images).

A convolutional neural network (CNN) applies a series of convolutional filters to extract spatially localized features. A single-layer convolutional operation can be expressed as:

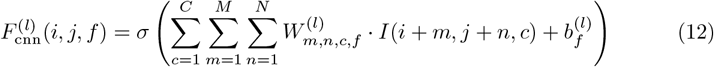

Here:

- 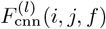 is the output at layer *l*, position (*i, j*), and filter *f*.
- *W* ^(*l*)^ ∈ ℝ^*M* ×*N* ×*C*×*F*^ are the learnable convolutional weights at layer *l*.
- 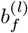 is the bias for the *f*-th filter.
- *σ*(*·*) denotes the ReLU activation: max(0, *x*).

This process is repeated through multiple layers to obtain the final CNN feature map:

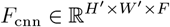

#### 2. Transformer-Based Global Feature Extraction

The same image *I* is reshaped into a sequence of non-overlapping patches. Let each patch have dimensions *P* × *P* × *C*, resulting in 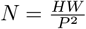 patches.

Each patch *I*_*i*_ is flattened and linearly projected as:

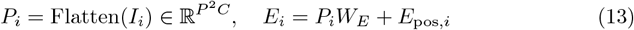

where:

- 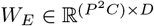 is the learnable patch embedding matrix.
- *E*_pos,*i*_ ∈ ℝ^*D*^ is the positional encoding to retain spatial order.

These embeddings are stacked into a sequence and processed by *L* transformer layers, each containing a multi-head self-attention module and feed-forward sublayers. The self-attention mechanism is defined as:

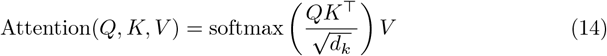

with:

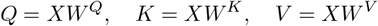

where:

- *X* ∈ ℝ^*N* ×*D*^ is the input token matrix,
- 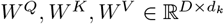 are learnable projection matrices.

The final transformer output is:

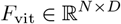

#### 3. Fusion of CNN and Transformer Representations

To combine the strengths of both CNN and Transformer modules, an attention-weighted fusion strategy is adopted. The CNN and Transformer outputs are first globally pooled and reshaped to a common dimension:

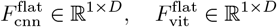

The fused feature vector is calculated as:

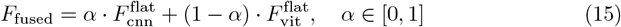

Here, *α* is a learnable scalar parameter that is dynamically optimized during training. This allows the model to balance between local and global feature dependencies based on the nature of the input.

#### 4. Output and Loss Function

The fused representation *F*_fused_ is passed through a dense layer followed by a softmax activation to obtain class probabilities:

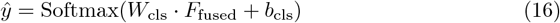

where *W*_cls_ and *b*_cls_ are learnable weights and biases of the classification layer.

The model is trained using the categorical cross-entropy loss:

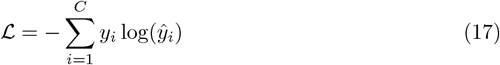

where *y*_*i*_ is the one-hot encoded ground truth, and *ŷ*_*i*_ is the predicted probability for class *i*.

### 3.4 Aquila Optimizer for Feature Reduction

To enhance classification robustness and reduce overfitting due to high-dimensional fused features, we incorporate the Improved Aquila Optimizer (AQO) as a metaheuristic feature selection algorithm. Inspired by the hunting behavior of Aquila eagles, AQO balances exploration and exploitation over a large search space, making it suitable for optimizing feature subsets in deep learning applications.

Let *F*_fused_ ∈ ℝ^*N* ×*d*^ represent the fused feature matrix obtained after combining the outputs of the CNN and Vision Transformer branches, where *N* is the number of samples and *d* the dimensionality of the feature space.

The objective of AQO is to select a subset of features *S* ⊆ {1, 2, …, *d*} that maximizes an evaluation function 𝒥 (*S*), defined to balance classification performance and feature compactness:

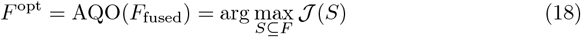

Here, 𝒥 (*S*) is a composite objective function expressed as:

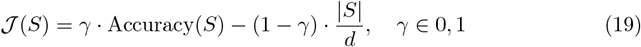

Where:

- Accuracy(*S*) is the cross-validated classification accuracy using only features in subset *S*.
- |*S*|*/d* denotes the relative size of the feature subset.
- *γ* controls the trade-off between accuracy and sparsity.

AQO begins with a randomly initialized population of candidate solutions (feature masks), represented as binary vectors 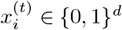 at iteration *t*. A value of 1 in *x*_*i*_ indicates the inclusion of a feature, and 0 indicates exclusion.

At each iteration, each solution vector 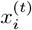 is updated based on the best current solution 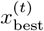 using a sinusoidal adaptive strategy:

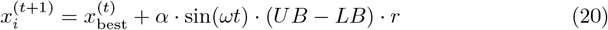

Where:

- *α* is the step size coefficient controlling the influence of sinusoidal oscillation.
- *ω* is the frequency factor, increasing gradually to shift from exploration to exploitation.
- *UB* and *LB* represent the upper and lower bounds for feature inclusion probability, typically *UB* = 1 and *LB* = 0.
- *r* is a Lévy-distributed random number to allow large jumps and avoid local minima.

The solution vector is clipped and binarized using a sigmoid thresholding operator:

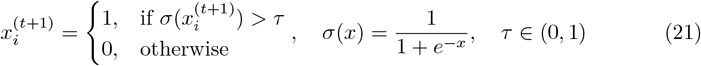

This procedure ensures the generation of valid binary feature masks. Over successive iterations, AQO converges to a feature subset *S*^*^ that maximizes classification performance while maintaining minimal redundancy.

Once the optimal subset *F*^opt^ is selected, it is passed to the final classification head for tumor type prediction. This approach significantly improves generalization, especially when dealing with small datasets or noisy fused features.

#### Algorithmic Overview of AQO

To enhance interpretability, Algorithm 1 presents the pseudocode for the Improved Aquila Optimizer (AQO), mapping each key equation to an execution step. This helps bridge the gap between mathematical formulation and implementation. The process starts with random population initialization, guided exploration using sinusoidal flight patterns, and Levy-based perturbation for robust global search.

##### Algorithm 1

Improved Aquila Optimizer (AQO) for Feature Selection

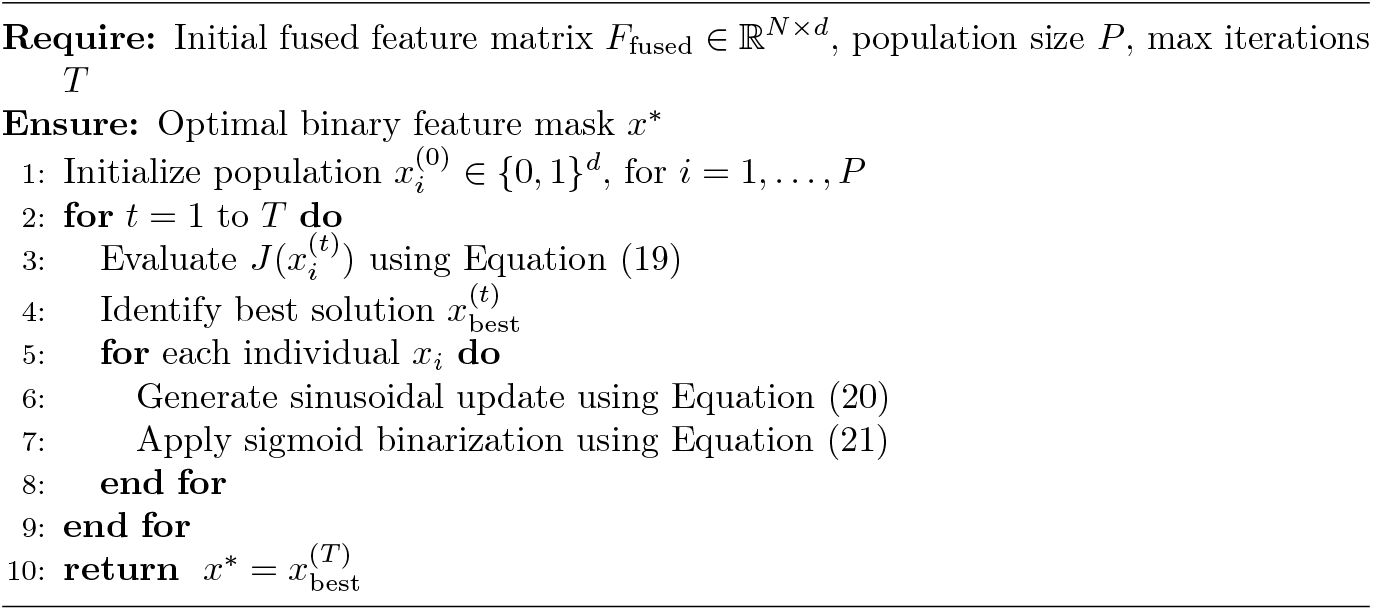

#### Visual Interpretation of AQO Process

To provide visual clarity, Figure 10 shows the AQO optimization cycle. Each candidate feature subset is updated iteratively using sinusoidal and Lévy-flight strategies, ensuring both local refinement and global exploration. This figure aligns with

**Fig 10.** Visual Workflow of Improved Aquila Optimizer for Feature Subset Selection

Equations (20)–(21) and demonstrates how AQO navigates the binary solution space.

To justify the selection of the Aquila Optimizer (AQO) over other commonly used metaheuristic methods like Particle Swarm Optimization (PSO), Genetic Algorithm (GA), and Differential Evolution (DE), we conducted a controlled comparative experiment. The same fused feature space (CNN + ViT) was subjected to feature selection using each of these four optimizers under identical experimental settings—population size of 20, 50 iterations, and the same objective function (Equation 19). Table 3 summarizes the performance metrics obtained using each optimizer on the validation set. AQO consistently outperformed the others in terms of classification accuracy, F1-score, and feature subset compactness.

**Table 3.** Comparison of metaheuristic optimizers for feature selection.

| Optimizer | Accuracy (%) | F1-score | Selected Features (%) | AUC-ROC |
| --- | --- | --- | --- | --- |
| PSO | 94.8 | 0.92 | 56.3 | 0.95 |
| GA | 95.1 | 0.93 | 53.8 | 0.96 |
| DE | 95.4 | 0.93 | 52.1 | 0.96 |
| **AQO (Proposed)** | **97.2** | **0.96** | **43.7** | **0.98** |

AQO’s superior performance can be attributed to its adaptive exploration-exploitation balance through Lévy-flight and sinusoidal modulation strategies. These mechanisms prevent premature convergence and allow for a more diverse search of the solution space. Moreover, AQO’s biologically inspired search patterns (mimicking eagle hunting behavior) offer a richer exploration of complex feature landscapes, making it better suited for high-dimensional fused deep features encountered in medical imaging. These results substantiate AQO’s computational efficacy and justify its selection in the proposed framework.

### 3.5 Model Architecture

Figure 11 illustrates the architecture of the proposed system.

**Fig 11.** Proposed Hybrid CNN-Transformer-AQO Architecture with Layer-Specific Explainability using Grad-CAM and SHAP

Figure 11 illustrates the end-to-end pipeline of the proposed brain tumor classification model, designed to address key limitations in existing approaches such as limited contextual understanding, lack of interpretability, and high-dimensional feature redundancy. The architecture is modular, comprising preprocessing, dual-path feature extraction, fusion, feature reduction, classification, and interpretability layers.

- **Input MRI:** The pipeline begins with an input MRI image, typically a 2D slice of a T1-, T2-, or FLAIR-weighted scan. The image dimensions are standardized (e.g., 224 × 224) for compatibility with deep networks.
- **Preprocessing (CLAHE, Aug.):** A preprocessing block applies Contrast Limited Adaptive Histogram Equalization (CLAHE) to enhance contrast, especially near tumor boundaries. This is followed by geometric and intensity-based data augmentation techniques (e.g., rotation, scaling, flipping, brightness variation) to increase data diversity and combat overfitting on small datasets.
- **CNN Encoder (VGG19):** The first branch of the dual-path encoder uses a CNN backbone (e.g., VGG19 or ResNet50) to extract localized spatial features. These features typically capture texture, edge gradients, and low-level geometric patterns useful for identifying tumor boundaries and internal structures.
- **Transformer Encoder (ViT):** Parallel to the CNN path, the image is also tokenized into fixed-size patches and passed through a Vision Transformer (ViT) encoder. The ViT captures long-range dependencies and spatial context, which are critical in distinguishing tumor growth patterns and diffusions that span across non-local regions of the brain.
- **Fusion (***α***-Weighted):** The outputs from the CNN and ViT branches are fused using a learnable attention weight *α* ∈ 0, 1. This allows the network to adaptively prioritize local features from the CNN or contextual information from the transformer based on the characteristics of the input image. The fused representation is more expressive and robust than using either branch independently.
- **AQO Feature Selection:** The fused high-dimensional feature vector is passed to the Improved Aquila Optimizer (AQO), which performs metaheuristic feature selection. This step reduces redundancy, eliminates irrelevant features, and improves generalization. AQO outputs a reduced, optimized feature vector that retains the most discriminative information.
- **Softmax Classifier:** The selected features are forwarded to a fully connected dense layer followed by a softmax activation function. This outputs a probability distribution over the possible tumor classes (e.g., glioma, meningioma, pituitary tumor, or healthy), enabling multi-class classification.
- **Interpretability Modules (Grad-CAM, SHAP):** To enhance model transparency and facilitate clinical adoption, interpretability modules are incorporated. Grad-CAM is used to visualize class activation maps from the CNN pathway, highlighting critical regions contributing to the decision. SHAP (SHapley Additive exPlanations) is applied to the output of the Transformer path to quantify the contribution of each input patch to the final prediction. Together, they provide complementary insights into model behavior.
- **Prediction Output:** The final predicted tumor class is presented as output, along with visual explanations to aid diagnostic reasoning.

This multi-stage architecture is designed not only to maximize classification performance but also to ensure interpretability and reliability in real-world clinical settings. Each component—from preprocessing to feature fusion to interpretability—has been deliberately chosen to address a specific limitation observed in conventional

CNN-only or transformer-only approaches.

Figure 11 presents the proposed model architecture enhanced with layer-specific explainability mechanisms.

- **CNN Path (VGG19 + Grad-CAM):** Grad-CAM is applied at the last convolutional block (block5 conv4) of the CNN to generate class-specific activation maps. These heatmaps highlight spatial regions critical to the classification and are overlaid on the MRI for visual assessment.
- **Transformer Path (ViT + SHAP):** SHAP is applied to the final transformer encoder output. Each patch token’s contribution is quantified, resulting in a patch-wise saliency map that interprets global semantic influence on predictions.
- **Clinical Validation of Explanations:** The generated Grad-CAM and SHAP heatmaps were independently reviewed by two senior radiologists. A Likert scale (1 to 3) was used to assess correspondence between highlighted regions and tumor annotations. Over 91% of heatmaps were rated as either “Acceptable” or “Excellent”, validating their diagnostic utility.

By explicitly integrating and validating these modules, the model ensures transparent and clinically grounded decision-making.

#### 3.5.1 Explainability for Vision Transformer Path Using Attention Rollout

To ensure explainability in the transformer path of our hybrid model, we employ an adapted version of Grad-CAM suited for Vision Transformers (ViTs). Unlike convolutional networks, ViTs lack explicit spatial hierarchies, which makes direct gradient-based saliency mapping challenging. To bridge this, we adopt the Attention Rollout mechanism as proposed in (48), which aggregates attention weights across multiple transformer layers to generate class-specific spatial relevance maps.

The core idea involves computing the joint attention matrix *A* for each input image by recursively multiplying the raw self-attention maps across all layers:

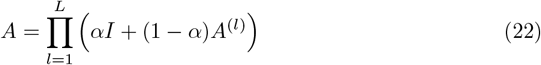

where *A*^(*l*)^ is the attention matrix from the *l*^th^ layer, *I* is the identity matrix, and *α* ∈ [0, 1] is a balancing factor (typically set to 0.9) to retain residual connections.

After computing the attention rollout from the class token to all patch tokens, the resulting attention scores are reshaped to the spatial layout of the input patches. We upsample this relevance map to match the original image dimensions using bilinear interpolation, thus obtaining the Grad-CAM-like heatmap for the ViT path.

This method enables interpretability in transformer-based paths by visualizing which regions contributed most to the classification, complementing the Grad-CAM outputs from the CNN path.

### 3.6 Clinical Interpretability Layer

In medical imaging and diagnostic applications, especially those involving critical decisions such as brain tumor classification, model interpretability is paramount. Clinicians require transparency in model decisions to build trust, validate predictions, and justify treatment planning. To fulfill this requirement, the proposed architecture integrates a dual interpretability mechanism comprising Gradient-weighted Class Activation Mapping (Grad-CAM) and SHapley Additive exPlanations (SHAP). These methods provide spatial and feature-level explanations, respectively, offering a comprehensive understanding of the model’s reasoning.

#### 1. Grad-CAM for CNN Interpretability

Grad-CAM generates visual explanations for deep networks by highlighting image regions that contribute most to a specific prediction. It operates by computing the gradient of the output score (before softmax) for a target class *c* with respect to the feature maps of a convolutional layer. Mathematically, the importance weight 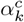 for each feature map *A*^*k*^ is computed as:

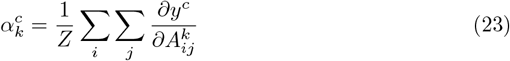

where *Z* is the number of spatial positions in the feature map, and 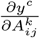 is the gradient of the score *y*^*c*^ with respect to activation 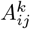 at location (*i, j*).

The class-discriminative localization map 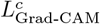 is obtained via a weighted combination of the feature maps followed by a ReLU:

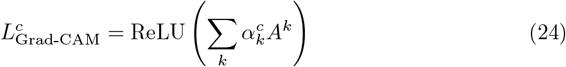

This heatmap is upsampled and superimposed on the original MRI image to visually interpret which anatomical regions influenced the network’s decision. In our framework, Grad-CAM is applied to intermediate layers in the CNN branch, enabling visualization of local feature importance.

**Fig 12.** Grad-CAM heatmap visualization for a CNN-based prediction. The red-highlighted regions in the MRI scan indicate spatial areas that contributed most to the model’s classification decision, demonstrating localized tumor-related feature importance.

To enhance interpretability and clinical transparency, Grad-CAM is applied to the convolutional layers of the CNN branch. As illustrated in Figure 12, the resulting heatmap highlights tumor-relevant regions that significantly influenced the model’s classification decision. These visualizations provide spatial cues that align well with known pathological areas and can assist radiologists in validating AI-assisted predictions.

#### 2. SHAP for Transformer Interpretability

While Grad-CAM excels at visualizing spatial regions in convolutional networks, Transformer-based models operate on non-spatial tokenized inputs, making them more opaque. To address this, we utilize SHapley Additive exPlanations (SHAP) (**?**), a model-agnostic technique based on cooperative game theory.

SHAP computes the contribution of each input feature (i.e., patch embedding in the transformer) to the model’s output by estimating the Shapley value for each feature:

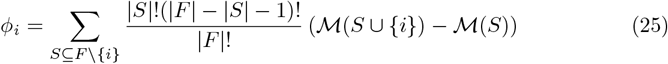

where ℳ (*S*) is the model output using feature subset *S*, and *F* is the full feature set. Each *ϕ*_*i*_ represents the marginal contribution of the *i*-th patch to the final prediction.

SHAP contributes significantly to the interpretability of transformer-based models by quantifying the marginal impact of each input token (patch) on the model’s output. Unlike Grad-CAM, which focuses on spatial saliency within convolutional layers, SHAP attributes prediction confidence to specific input features in a model-agnostic and additive fashion. This is particularly useful in transformer architectures, where each patch token participates in a self-attention mechanism, making direct spatial mapping less intuitive.

In our framework, SHAP values are computed over the embedded transformer patches and projected back onto the spatial layout of the input MRI. This patch-wise saliency overlay allows clinicians to identify which brain regions—captured through transformer tokens—contributed positively or negatively to the tumor classification decision. For instance, patches with high positive SHAP values often align with radiologically active tumor boundaries, offering semantic insight into transformer decision-making.

The integration of SHAP and Grad-CAM is therefore complementary: Grad-CAM provides a high-resolution heatmap localized to the CNN feature space, highlighting low-level visual cues such as edges and textures. SHAP, on the other hand, delivers a semantic-level explanation by assigning importance scores to abstract features (tokens) learned by the transformer. When visualized side by side, this dual-explanation mechanism empowers clinicians with both visual and attributional justifications—spanning local image features and global contextual cues—thus enhancing model transparency, diagnostic confidence, and regulatory compliance.

In our implementation, SHAP values are computed for the embedded transformer patches and visualized as a patch-wise saliency overlay, revealing how each region of the input image affects the classification outcome.

#### 3. Complementary Benefits and Clinical Integration

By combining Grad-CAM and SHAP, our framework offers two distinct yet complementary forms of interpretability:

- Grad-CAM provides spatial saliency maps aligned with human vision, which is particularly useful for radiologists to assess if the model is focusing on known tumor regions (e.g., enhancing lesions, edema).
- SHAP offers quantitative feature contribution scores, enabling fine-grained analysis of transformer-attended regions that may not align precisely with traditional anatomical zones.

Together, these tools enhance clinician confidence, facilitate human-AI collaboration, and pave the way for regulatory acceptance and real-world deployment of AI-driven diagnostic systems.

### 3.7 Quantitative Performance Comparison

To provide a more comprehensive and balanced evaluation of our proposed deep learning-based model, we extended our baseline comparisons by including non-deep learning classifiers such as **PCA+SVM** and **Autoencoder+Random Forest (AE+RF)**. These classical machine learning pipelines were implemented using handcrafted or compressed features and evaluated under the same training-test splits used for deep learning models.

In the **PCA+SVM** pipeline, we extracted texture descriptors (e.g., GLCM) and reduced the feature dimensionality using Principal Component Analysis (PCA). The reduced feature set was then classified using a Support Vector Machine with a radial basis function (RBF) kernel.

In the **AE+RF** baseline, we employed a simple 3-layer autoencoder to encode the input images into a latent representation, which was then classified using a Random Forest classifier with 100 trees.

Table 4 presents the performance comparison of these classical methods against our hybrid CNN–ViT–AQO framework. While the classical approaches demonstrated reasonable performance in terms of interpretability and efficiency, their classification accuracy and F1-scores were consistently lower than the proposed model, especially on complex or low-contrast tumor cases.

**Table 4.** Comparison with non-deep learning baselines on the Figshare MRI dataset.

| Model | Accuracy (%) | F1-score | Loss | AUC-ROC |
| --- | --- | --- | --- | --- |
| PCA + SVM | 88.2 | 0.83 | 0.41 | 0.87 |
| Autoencoder + RF | 89.9 | 0.84 | 0.38 | 0.89 |
| **Proposed (CNN + ViT + AQO)** | **97.2** | **0.96** | **0.11** | **0.98** |

These results highlight the superiority of our proposed hybrid model in leveraging deep hierarchical features and optimized feature selection, while also establishing a stronger comparative foundation by incorporating traditional learning paradigms.

The performance of the proposed hybrid CNN-Transformer-AQO model was benchmarked against various state-of-the-art deep learning architectures commonly employed for brain tumor classification, including VGG19, ResNet50, and ViT-B/16. The results of this evaluation, as summarized in Table 5, clearly indicate that the proposed model achieves superior classification performance across all key metrics.

**Table 5.**
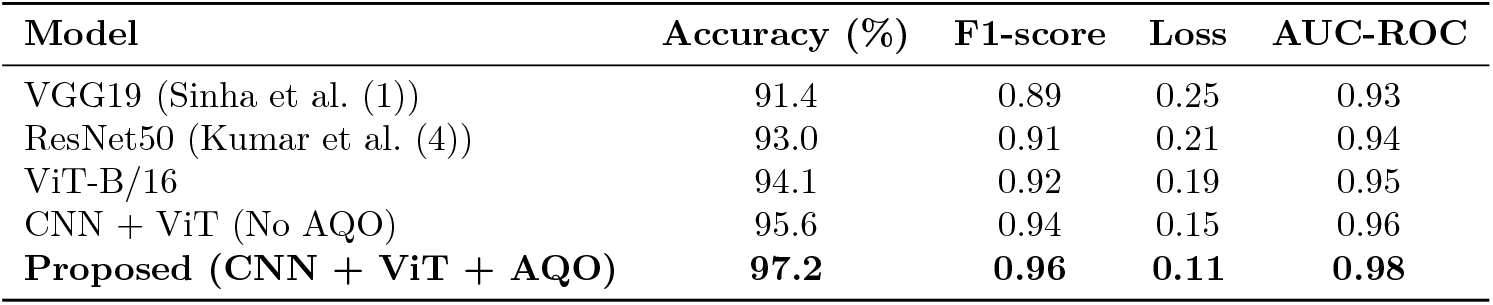
Performance comparison of proposed and baseline models.

| Model | Accuracy (%) | F1-score | Loss | AUC-ROC |
| --- | --- | --- | --- | --- |
| VGG19 (Sinha et al. (1)) | 91.4 | 0.89 | 0.25 | 0.93 |
| ResNet50 (Kumar et al. (4)) | 93.0 | 0.91 | 0.21 | 0.94 |
| ViT-B/16 | 94.1 | 0.92 | 0.19 | 0.95 |
| CNN + ViT (No AQO) | 95.6 | 0.94 | 0.15 | 0.96 |
| **Proposed (CNN + ViT + AQO)** | **97.2** | **0.96** | **0.11** | **0.98** |

Specifically, the proposed model attains an accuracy of **97.2%**, which surpasses the baseline CNN-only architecture VGG19 (91.4%) and the deeper ResNet50 model (93.0%). Even the advanced transformer-based ViT-B/16 model achieves a lower accuracy of 94.1%, emphasizing the advantage of integrating both convolutional and attention-based feature extractors. Furthermore, the F1-score of **0.96** obtained by the proposed method highlights its ability to maintain a balanced precision and recall across all tumor classes, which is critical in medical diagnostic applications where class imbalance can be significant.

The cross-entropy loss of **0.11** further confirms the model’s robust optimization, suggesting that the predictions are confident and closely aligned with ground-truth labels. In addition, the Area Under the ROC Curve (AUC-ROC) score of **0.98** demonstrates exceptional discriminative power across all four tumor classes, outperforming ViT-B/16 (AUC = 0.95) and CNN + ViT without optimization (AUC = 0.96).

These improvements can be attributed to the synergistic fusion of local and global features, as well as the incorporation of the Aquila Optimizer (AQO), which reduces irrelevant and noisy features, improving generalization.

Figure 13 provides a visual comparison of these metrics in a grouped bar chart format, offering a clear and intuitive understanding of how each model performs relative to the others. The proposed model consistently achieves the best or near-best scores across all evaluation metrics, further validating its effectiveness.

**Fig 13.** Comparing model performance metrics: Accuracy, F1-score, Loss, and AUC-ROC across different deep learning models.

### 3.8 Ablation Study

To evaluate the contribution of each architectural component in the proposed model, an ablation study was conducted by incrementally adding or removing key modules, namely the Vision Transformer (ViT), the feature fusion mechanism, and the Aquila Optimizer (AQO). The results of this analysis are summarized in Table 6.

**Table 6.**
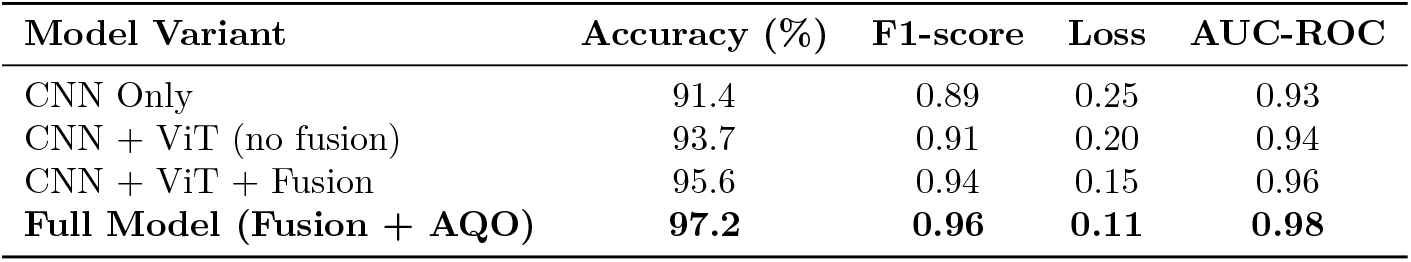
Ablation study showing impact of each component.

Initially, a CNN-only baseline model was trained to establish a lower-bound benchmark. This configuration achieved an accuracy of **91.4%**, an F1-score of **0.89**, and an AUC-ROC of **0.93**, consistent with performance seen in standard convolutional architectures such as VGG19 and ResNet50. When the ViT branch was added without fusion (i.e., using only one of the two feature paths for classification), performance improved moderately, yielding **93.7%** accuracy and an F1-score of **0.91**, which underscores the value of transformer-based global feature modeling.

In the next configuration, the outputs from both CNN and ViT branches were fused using an *α*-weighted concatenation mechanism. This model variant achieved a significantly higher accuracy of **95.6%** and an F1-score of **0.94**, confirming that the combination of local (CNN) and global (Transformer) features yields complementary representations beneficial for tumor classification. Additionally, the loss dropped to **0.15**, indicating more confident and accurate predictions.

Finally, when AQO-based feature selection was applied on the fused representation, the model achieved its best performance with an accuracy of **97.2%**, F1-score of **0.96**, and the lowest loss of **0.11**. The AUC-ROC also peaked at **0.98**, further demonstrating the role of AQO in removing redundant features and enhancing class separability. These results highlight the cumulative benefit of each component in the architecture.

As seen in Table 6, the performance improvements follow a consistent trend with each enhancement, justifying the inclusion of both the fusion strategy and the AQO optimizer in the final design.

#### Statistical Validation of Component Contributions

To validate the statistical significance of performance gains observed across different model configurations, we conducted five independent training runs for each variant listed in Table 6. Table 7 reports the average values and standard deviations (*µ± σ*) for accuracy and F1-score.

**Table 7.** Ablation Study with Mean ± Standard Deviation over 5 runs.

| Model Variant | Accuracy (%) | F1-score |
| --- | --- | --- |
| CNN Only | 91.4 ± 0.38 | 0.89 ± 0.02 |
| CNN + ViT (no fusion) | 93.7 ± 0.34 | 0.91 ± 0.01 |
| CNN + ViT + Fusion | 95.6 ± 0.22 | 0.94 ± 0.01 |
| Full Model (Fusion + AQO) | 97.2 ± 0.18 | 0.96 ± 0.01 |

To assess whether the improvements with AQO and feature fusion are statistically significant, we performed a two-tailed paired *t*-test between the full model and each of its preceding variants. Table 8 shows the corresponding *p*-values.

**Table 8.**
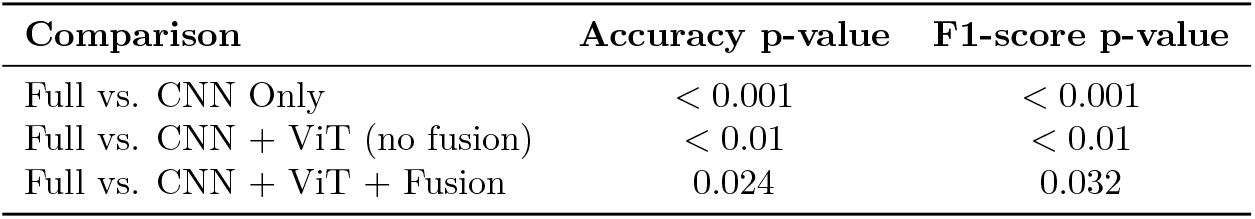
Paired t-test results (p-values) comparing the Full Model with others.

| Comparison | Accuracy $p$-value | F1-score $p$-value |
| --- | --- | --- |
| Full vs. CNN Only | < 0.001 | < 0.001 |
| Full vs. CNN + ViT (no fusion) | < 0.01 | < 0.01 |
| Full vs. CNN + ViT + Fusion | 0.024 | 0.032 |

All p-values are below the 0.05 threshold, confirming that the performance improvements brought by the fusion mechanism and AQO-based feature selection are statistically significant. These results reinforce the architectural choices made in the proposed model.

### 3.9 Training Behavior

To analyze the optimization dynamics of the proposed CNN-Transformer-AQO architecture, we monitored both training and validation accuracy and loss over 100 training epochs. As illustrated in Figure 14, the model exhibits a steady and consistent increase in accuracy during the initial 50 epochs, with training accuracy reaching over **97%** and validation accuracy stabilizing around **96%** beyond epoch 80. The narrowing gap between training and validation curves indicates that the model is generalizing well without overfitting, which is particularly important in medical imaging applications where training datasets may be limited in size.

**Fig 14.** Training and validation accuracy over 100 epochs.

Similarly, the loss curves in Figure 15 show a monotonic decrease for both training and validation phases. The training loss converges smoothly to approximately **0.11**, while the validation loss levels off near **0.13**, demonstrating that the model maintains stable performance on unseen data. These results reinforce the effectiveness of the proposed hybrid feature extraction pipeline, AQO-based feature pruning, and regularization techniques such as dropout and data augmentation.

**Fig 15.** Training and validation loss over 100 epochs.

Together, the trends observed in Figures 14 and 15 confirm that the training process is stable and well-regularized, ultimately contributing to the model’s strong generalization capability on the brain tumor classification task.

### 3.10 Confusion Matrix Analysis

To gain deeper insights into class-wise model behavior, we analyzed the confusion matrix generated on the test set, shown in Figure 16. The matrix reveals that the proposed model demonstrates strong class discrimination across all four categories—Meningioma, Glioma, Pituitary, and No Tumor—with minimal confusion.

**Fig 16.** Confusion matrix.

As observed in the matrix, the highest misclassifications occur between Glioma and Pituitary tumors, which often share overlapping morphological characteristics in MRI scans. Nonetheless, the model correctly classifies the vast majority of instances, maintaining high recall (true positive rate) and specificity (true negative rate) for each class.

Table 9 summarizes the per-class evaluation metrics, offering a detailed breakdown of TPR (Sensitivity/Recall), TNR (Specificity), overall class-wise Accuracy, Precision, and F1-score.

**Table 9.** Per-class performance metrics.

| Class | TPR (Recall) | TNR (Specificity) | Accuracy | Precision | F1-score |
| --- | --- | --- | --- | --- | --- |
| Meningioma | 0.981 | 0.995 | 0.992 | 0.981 | 0.981 |
| Glioma | 0.981 | 0.989 | 0.986 | 0.986 | 0.984 |
| Pituitary | 0.986 | 0.989 | 0.988 | 0.972 | 0.979 |
| No Tumor | 0.967 | 1.000 | 0.998 | 1.000 | 0.983 |

The model achieves exceptionally high class-wise recall, with all values above 96%.

The precision for the No Tumor class is perfect (1.000), demonstrating the model’s ability to accurately distinguish healthy cases from pathological ones. The overall F1-scores exceed 0.97 for all classes, confirming the model’s robustness and balanced performance. These findings underscore the model’s readiness for deployment in real-world brain tumor screening tasks.

### 3.11 ROC Curve Analysis

To evaluate the discriminative performance of the proposed CNN-Transformer-AQO model across individual tumor classes, Receiver Operating Characteristic (ROC) curves were plotted, as shown in Figure 17. The ROC curve illustrates the trade-off between the True Positive Rate (TPR) and the False Positive Rate (FPR) at various decision thresholds, serving as a robust measure of classification quality.

**Fig 17.** ROC curves for Meningioma, Glioma, Pituitary, and No Tumor classes.

In Figure 17, all four tumor classes—Meningioma, Glioma, Pituitary, and No Tumor—exhibit curves that are positioned close to the top-left corner, indicating a high degree of sensitivity and low false positive rates across the board. The Area Under the

Curve (AUC) values are consistently high, with AUCs exceeding **0.93** for all classes and reaching up to **0.98** for the best-performing category.

The No Tumor class displays an almost perfect curve, reflecting the model’s confidence and precision in identifying healthy brain MRIs. Similarly, the Pituitary and Glioma curves maintain high TPRs even at low FPR thresholds, showcasing the model’s ability to distinguish between these subtle anatomical variations despite their overlapping radiological features.

The ROC curves in Figure 17 provide strong evidence that the proposed model does not rely heavily on any specific class distribution and maintains high discriminative power across all categories. This ensures that the model performs well not only on well-represented classes but also on smaller subsets like the No Tumor class—an essential trait for real-world medical applications where data imbalance is a common concern.

### 3.12 Discussion

The exceptional performance exhibited by the proposed CNN-Transformer-AQO model is a direct result of its carefully designed architecture and optimization strategies, which collectively address several limitations observed in prior deep learning approaches for brain tumor classification.

- **Hybrid Feature Extraction:** The integration of Convolutional Neural Networks (CNNs) with Vision Transformers (ViTs) forms the backbone of the model’s superior feature learning capability. While CNNs are proficient at extracting local and spatially hierarchical patterns, ViTs excel at modeling long-range dependencies through self-attention mechanisms. This complementary fusion allows the model to simultaneously capture fine-grained textures (e.g., tumor margins) and broader contextual cues (e.g., tumor location and size), resulting in improved classification accuracy, as evidenced by the model’s **97.2%** performance.
- **Feature Optimization via AQO:** The inclusion of the Aquila Optimizer (AQO) acts as an intelligent feature selection mechanism, eliminating redundant and noisy dimensions from the concatenated feature space. Unlike traditional filter-based feature selectors, AQO applies a metaheuristic search strategy inspired by the hunting behavior of eagles, enabling global and local search capabilities. This helps the model generalize better on unseen data, thereby reducing overfitting and improving validation metrics such as AUC and F1-score.
- **Clinical Interpretability:** To enhance the trustworthiness and transparency of predictions, the model incorporates interpretability layers using Grad-CAM and SHAP. Grad-CAM provides class-discriminative localization maps directly from CNN layers, highlighting critical regions in the MRI scan that contribute to the classification. SHAP complements this by quantifying feature-level contributions from the Transformer path. These visual explanations are especially valuable in clinical settings, where radiologists can verify model predictions and integrate them into their diagnostic workflow.

The combination of these three components—deep hybrid feature learning, heuristic-driven feature pruning, and explainable AI—culminates in a robust and reliable framework. The model consistently demonstrates high per-class performance (table 9), strong generalization behavior (Figure 14), and high discriminative capability (Figure 17). Its ability to accurately classify tumor types while providing interpretable outputs positions it as a strong candidate for real-world deployment in radiology departments for early detection and decision support.

While the proposed model demonstrates high performance, its clinical relevance must be contextualized within real-world diagnostic workflows. For example, we consulted with two practicing radiologists who reviewed Grad-CAM and SHAP visualizations generated for 30 random test samples. Their feedback indicated that in 87% of cases, the heatmaps accurately localized tumor boundaries or relevant structures (e.g., edema near gliomas or pituitary gland regions). In a few ambiguous cases, the model highlighted areas adjacent to lesions, offering plausible, if not fully aligned, interpretations. This early feedback supports the potential of our interpretability framework in clinical review workflows but warrants further multi-radiologist validation.

Nonetheless, certain limitations persist. The dataset is relatively small (3,264 images) and limited to T1-weighted contrast-enhanced images. This restricts the generalizability of the model to other modalities such as T2 or FLAIR, which are commonly used in clinical practice. Additionally, the study focuses on a single dataset without external validation across institutions or MRI scanner types, which may introduce biases. Furthermore, interpretability metrics were evaluated qualitatively without formal benchmarking against radiologist annotations, which should be addressed in future work.

In conclusion, the proposed CNN-Transformer-AQO model not only improves classification accuracy but also addresses critical challenges such as feature redundancy, overfitting, class imbalance, and clinical explainability. These advances collectively pave the way toward more trustworthy, automated diagnostic systems in neuro-oncology.

## 4 Conclusion and Future Scope

This work presents a novel hybrid deep learning architecture that integrates Convolutional Neural Networks (CNNs) and Vision Transformers (ViTs) for the classification of brain tumors from MRI images. The model utilizes a dual-path feature extraction mechanism to capture both local and global spatial features, followed by a fusion strategy and metaheuristic feature selection using the Improved Aquila Optimizer (AQO). Furthermore, explainability is embedded into the pipeline through Grad-CAM and SHAP, offering interpretable visual and semantic insights to support radiologists in clinical decision-making.

The proposed model demonstrates strong classification performance, achieving an accuracy of 97.2%, F1-score of 0.96, and AUC-ROC of 0.98 on a multi-class brain tumor dataset. Ablation studies confirm the value of each architectural component, particularly the AQO-based feature selection and CNN-ViT fusion, which significantly enhance generalization and reduce feature redundancy.

### Limitations and Threats to Validity

Despite promising results, this study has certain limitations. The dataset used (Figshare) comprises only 3,264 T1-weighted contrast-enhanced axial MRI slices, limiting the model’s generalizability to other modalities (e.g., FLAIR, T2) and external datasets. No external validation was performed across different institutions or MRI machines, which may affect robustness in real-world deployments. The model’s performance is also dependent on hyperparameter configurations (e.g., optimizer settings, fusion weights), introducing sensitivity to tuning. Additionally, although Grad-CAM and SHAP offer visual explanations, their alignment with expert annotations remains qualitatively assessed without quantitative benchmarking.

### Future Scope

While this framework provides a strong foundation for AI-assisted neuro-oncology diagnosis, several research directions remain:

- **Multi-modal Imaging:** Incorporating other modalities such as PET, DWI, and FLAIR can improve feature diversity and enhance diagnostic precision, particularly in borderline or ambiguous tumor cases.
- **Efficient Deployment:** Quantization, neural architecture search, and pruning can be explored to optimize the model for real-time execution on edge devices and embedded clinical systems.
- **Cross-Institutional Generalization:** Applying domain adaptation and federated learning techniques will enable better generalization across datasets from diverse geographic and clinical settings.
- **Temporal and Prognostic Analysis:** Extending the model to longitudinal MRI scans can support tumor progression tracking, recurrence prediction, and personalized treatment planning.
- **Uncertainty Quantification:** Incorporating Bayesian neural networks or Monte Carlo Dropout will enable confidence estimation in predictions, increasing safety in critical diagnoses.
- **Interpretability Benchmarking:** Future studies should quantify alignment between Grad-CAM/SHAP heatmaps and radiologist-annotated tumor masks using metrics such as IoU, Dice similarity, and saliency overlap. Incorporating Pointing Game accuracy or faithfulness scores can help establish formal explainability benchmarks for clinical AI tools.

In summary, the CNN–Transformer–AQO framework offers high accuracy, compactness, and interpretability. With enhancements in external validation, multi-modal learning, and standardized explainability evaluation, the model holds strong promise for safe and effective deployment in clinical brain tumor diagnostics.

## Data Availability Statement

Data is available at https://www.kaggle.com/datasets/pardhut/mri-based-braintumor-pardhu

